# Surgical Diseases in North Korea: in view of North Korean medical journals

**DOI:** 10.1101/2020.10.25.20217141

**Authors:** Sejin Choi, Taehoon Kim, Soyoung Choi, Hee Young Shin

**Affiliations:** Department of Translational Medicine, Seoul National University College of Medicine, Seoul, Republic of Korea; Seoul Detention Center, Ministry of Justice, Seoul, Republic of Korea; Seoul National University College of Medicine, Seoul, Republic of Korea; Institute for Health and Unification Studies, Seoul National University College of Medicine, Seoul, Republic of Korea; Department of Pediatrics, Seoul National University College of Medicine, Seoul, Republic of Korea; The Republic of Korea National Red Cross, Seoul, Republic of Korea

**Keywords:** North Korea, surgery, research trends

## Abstract

**Introduction:** The demand for surgical diseases in North Korea is not fully reported despite its clinical significance. This study is to indirectly assess contemporary research trends and infrastructure status on surgical diseases in North Korea.

**Methods:** We analyzed articles of journal ‘*Surgery*’ published in Kim Jong-un era (2012-2018). The framework for categorization was primarily based on the disease entities, surgery specialty, and research methodology.

**Results:** 1,792 articles in 28 issues were included in the study. The frequency of detailed surgery cases and the characteristics of their specialty were investigated. The types of medical imaging techniques and anesthetics commonly utilized in clinical fields in North Korea were also evaluated.

**Conclusions:** An obvious disjunction between actual disease prevalence and research trends mirrors the lack of surgical technique and infrastructure. Further evaluation and attention to the surgical system in North Korea are required.

## INTRODUCTION

In general, information on the health and disease status of 24 million North Koreans is under veil due to the limited access to the country itself. (Spoorenberg and Schwekendiek 2012) Only small number of people have witnessed North Korean healthcare, and many of those experiences have been confined to Pyongyang, the capital city of North Korea. Few international organizations functioned as sources of data, but it has always been a challenge to gather reliable data. (Lee, Yoon et al. 2013) Also, a limited number of peer-reviewed articles written in English by North Korean medical researchers made it more difficult to capture the whole picture.

There are some facts known that the life expectancy of North Koreans is 71.69 years as of 2016 (Koen and Beom 2020) and that the number one cause of death is cardiovascular disease. (Lee, Yoon et al. 2013) However, details are still missing and incomplete. Also, reports have disproportionately focused more on communicable diseases (Park, Choi et al. 2018), including parasitic diseases (Lee, Seo et al. 2018, Chang 2019) and tuberculosis (Seung and Linton 2013, Seung, Franke et al. 2016) or nutritional issues in North Korea.

Recently, there have been efforts to grasp the reality by investigating refugees from North Korea instead of current residents of North Korea (Park, Kim et al. 2014) or through the North Korean medical journals in the fields of internal medicine (Kim, Ha et al. 2018, Ha and Lee 2019), pediatrics, obstetrics and gynecology (Ro, Kim et al. 2019) and psychiatry. (Kim and Jeon 2020) These attempts helped us understand the status quo of North Korea’s medical needs. For example, importance of non-communicable diseases in North Korea, which were underestimated got spotlights. (Ha and Lee 2019)

However, there are only few reports about the status of surgical diseases and surgical environments in North Korea (Park, Roh et al. 2015) while the surgery needs are clear, especially to reduce maternal-child mortality, to lighten the burden of trauma, and to treat life-threatening congenital anomalies as emphasized in WHO Global Initiative for Emergency and Essential Surgical Care.

According to Institute for Health Metrics and Evaluation, (http://www.healthdata.org/north-korea, accessed date: 2019-06-01 9:14 pm), the top 11 causes of death in North Korea are stroke, COPD, ischemic heart disease, liver cancer, lung cancer, Alzheimer’s disease, stomach cancer, road injuries, lower respiratory infect, hypertensive heart disease, and cirrhosis. Among these, 5 disease categories - hemorrhagic stroke, liver cancer, lung cancer, stomach cancer, road injuries-require surgeries to reduce mortality and increase survival chance.

Providing surgical treatment can be highly cost-effective, and having a clear vision on the burden of surgical diseases can help high-income countries to prioritize the usage of medical aids for North Korea. (Meara, Leather et al. 2015, Ng-Kamstra, Greenberg et al. 2016)

In this study, characteristics of medical researches on surgical diseases in North Korea were investigated through the North Korean medical journal “Surgery”. Also, this study aims to indirectly assess the demand for surgical diseases and status of infrastructure for surgeries –like anesthesia or radiological imaging in North Korea.

## METHODS

All articles published in North Korean Journal ‘Surgery [*Oegwa*]’ from 2012, when Kim Jong-un became the Chairman of the Workers’ Party of North Korea, to 2018 were reviewed. The journal issues were obtained from the Ministry of Unification’s ‘North Korea Resource Center’ in Seoul, South Korea. Articles were reviewed and analyzed by three reviewers – medical doctor, North Korea expert, and medical student (SJ, SY, TH)

### Variables

Disease entities were categorized into communicable, non-communicable, maternal, neonatal, nutritional, injury/trauma, and others according to the previous study. (Rose, Chang et al. 2014) Surgery specialty that covered each disease was classified into General Surgery; Cardiovascular & Thoracic Surgery; Obstetrics and Gynecology; Neurological surgery; Ophthalmic surgery; Oral maxillofacial surgery; Orthopedic surgery; Otorhinolaryngology; Urology; Plastic & Reconstructive Surgery; Rehabilitation Medicine; Anesthesiology according to the specialty category introduced by American College of Surgeons. (https://www.facs.org/education/resources/medical-students/faq/specialties)

Research methodology of each article was identified as one of the observational, randomized controlled trial (RCT), prospective cohort, retrospective cohort, case-control, case report or series basic science or experimental, systematic review, meta-analysis, Level 5 (Editorial, educational, letter, perspective), non-systematic review of literature, qualitative paper, mixed study, and other.

## RESULTS

### 1. Structure

In the North Korean journal ‘Surgery[Oegwa]’, an issue was published every quarter. Each issue had average of 64 articles inside. A total of 1,792 articles in 28 issues were published in the Journal ‘Surgery’ from 2012 to 2018. Articles were all written in Korean and could be classified into seven different types – Editorial, Original Article, Discussion, Experience, Review article, Case report or series, and Creative Invention. The distribution of article types is as follows in Table 1. About 60% of articles were original articles, and about 15% were case reports. “Creative invention” section was newly started in 2018.

**Table 1.** Distribution of article types.

|  | Original Article | Experience | Discussion | Review | Case | Editorial | Invention | Total |
| --- | --- | --- | --- | --- | --- | --- | --- | --- |
| **2012** | 127<br>(52.7%) | 31<br>(12.9%) | 26<br>(10.8%) | 23<br>(9.5%) | 34<br>(14.1%) | 0<br>(0.0%) | 0<br>(0.0%) | 241 |
| **2013** | 132<br>(54.8%) | 21<br>(8.7%) | 18<br>(7.5%) | 25<br>(10.4%) | 44<br>(18.3%) | 1<br>(0.4%) | 0<br>(0.0%) | 241 |
| **2014** | 133<br>(61.6%) | 14<br>(6.5%) | 16<br>(7.4%) | 16<br>(7.4%) | 34<br>(15.7%) | 3<br>(1.4%) | 0<br>(0.0%) | 216 |
| **2015** | 162<br>(67.5%) | 22<br>(9.2%) | 0<br>(0.0%) | 18<br>(7.5%) | 34<br>(14.2%) | 4<br>(1.7%) | 0<br>(0.0%) | 240 |
| **2016** | 165<br>(57.1%) | 26<br>(9.0%) | 24<br>(8.3%) | 26<br>(9.0%) | 44<br>(15.2%) | 4<br>(1.4%) | 0<br>(0.0%) | 289 |
| **2017** | 172<br>(62.1%) | 18<br>(6.5%) | 25<br>(9.0%) | 26<br>(9.4%) | 33<br>(11.9%) | 3<br>(1.1%) | 0<br>(0.0%) | 277 |
| **2018** | 168<br>(58.3%) | 17<br>(5.9%) | 28<br>(9.7%) | 25<br>(8.7%) | 43<br>(14.9%) | 4<br>(1.4%) | 3<br>(1.0%) | 288 |
| **Total** | 1059<br>(59.1%) | 149<br>(8.3%) | 137<br>(7.6%) | 159<br>(8.9%) | 266<br>(14.8%) | 19<br>(1.1%) | 3<br>(0.2%) | 1792 |

**Table 2.** Articles by disease category.

|  | Trauma | % | Neonatal /Congenital/Pediatric | % | communicable | % | Non-communicable | % | Basic Science | % |
| --- | --- | --- | --- | --- | --- | --- | --- | --- | --- | --- |
| **2012** | 53 | 22.0 | 14 | 5.8 | 3 | 1.2 | 79 | 32.8 | 20 | 8.3 |
| **2013** | 55 | 22.8 | 14 | 5.8 | 2 | 0.8 | 103 | 42.7 | 15 | 6.2 |
| **2014** | 54 | 25.0 | 8 | 3.7 | 1 | 0.5 | 85 | 39.4 | 21 | 9.7 |
| **2015** | 58 | 24.2 | 12 | 5.0 | 2 | 0.8 | 90 | 37.5 | 18 | 7.5 |
| **2016** | 71 | 24.6 | 9 | 3.1 | 1 | 0.3 | 108 | 37.4 | 20 | 6.9 |
| **2017** | 68 | 24.5 | 15 | 5.4 | 1 | 0.4 | 94 | 33.9 | 27 | 9.7 |
| **2018** | 101 | 35.1 | 10 | 3.5 | 0 | 0.0 | 72 | 25.0 | 22 | 7.6 |
| **Total** | 460 | 25.7 | 82 | 4.6 | 10 | 0.6 | 631 | 35.2 | 143 | 8.0 |

**Table 3.** Distribution of articles by specialty.

|  | NS | OS | PRS | CS | Uro | GS | A | Others | Total |
| --- | --- | --- | --- | --- | --- | --- | --- | --- | --- |
| **2012** | 16 | 74 | 17 | 25 | 12 | 79 | 12 | 6 | 241 |
| **2013** | 14 | 79 | 12 | 25 | 10 | 80 | 12 | 9 | 241 |
| **2014** | 9 | 61 | 13 | 23 | 13 | 56 | 14 | 27 | 216 |
| **2015** | 19 | 59 | 24 | 16 | 13 | 60 | 21 | 28 | 240 |
| **2016** | 18 | 85 | 27 | 26 | 17 | 79 | 9 | 28 | 289 |
| **2017** | 20 | 68 | 15 | 21 | 9 | 89 | 22 | 33 | 277 |
| **2018** | 15 | 105 | 27 | 19 | 5 | 86 | 19 | 12 | 288 |
| **Total** | 111 | 531 | 135 | 154 | 79 | 530 | 109 | 143 | 1792 |
| **%** | 6.2% | 29.6% | 7.5% | 8.6% | 4.4% | 29.5% | 6.1% | 8.0% | 100.0% |

Note. NS: Neurosurgery; OS: Orthopedic Surgery; PRS: Plastic and Reconstructive Surgery; CS: Cardiothoracic Surgery; Uro: Urology; GS: General Surgery; A: Anesthesiology

**Table 4.**
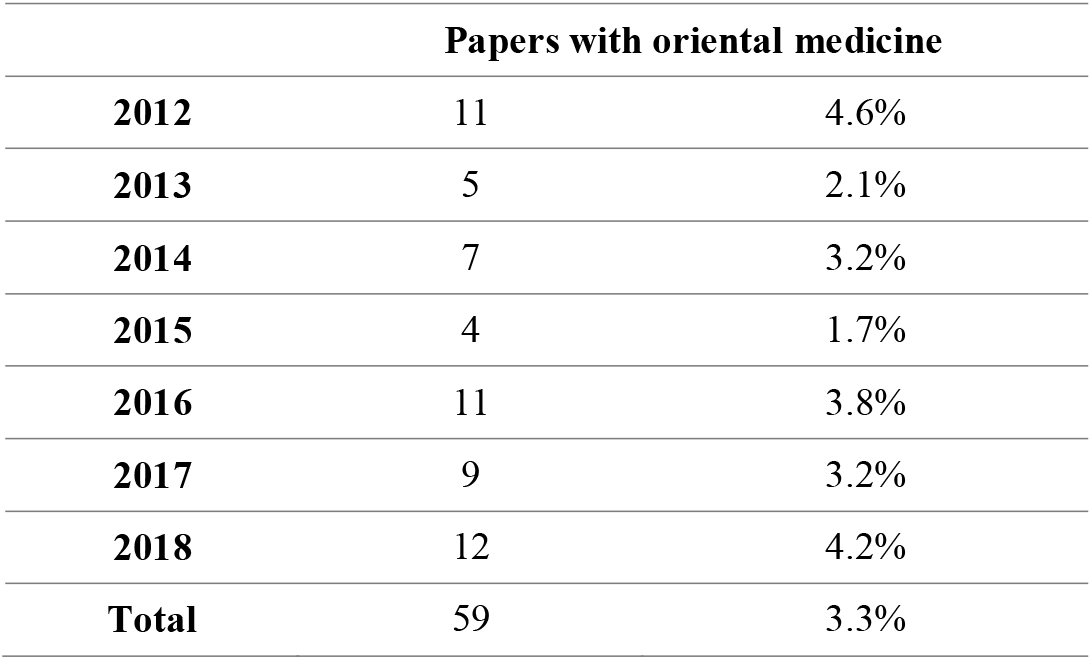
Papers that used oriental medicine for surgical diseases.

**Table 5.**
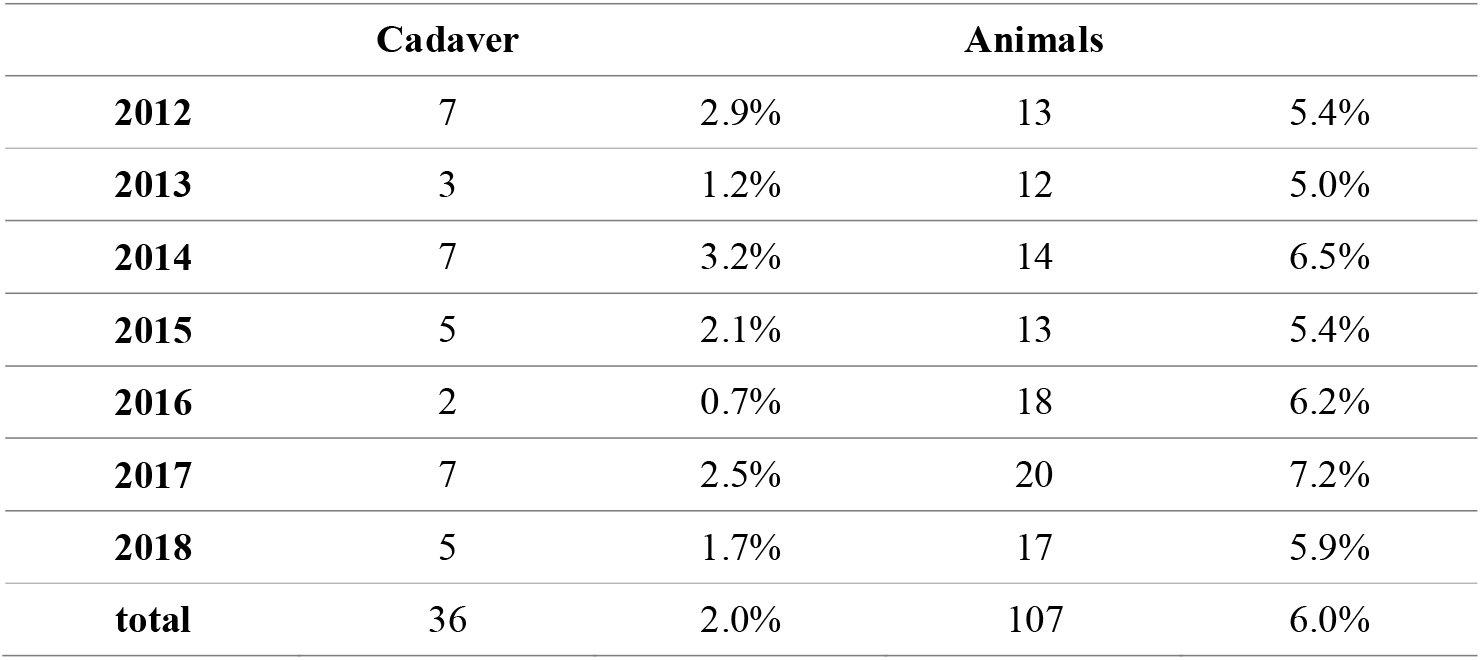
Studies using cadaver or animals.

**Table 6.1.**

|  | **Total articles** | **Reference of recent 5 years/total reference** | **Articles with reference** | **Articles without reference** | **Average number of references** | **Average number of references\*** |
| --- | --- | --- | --- | --- | --- | --- |
| **2012** | 241 | 25% | 145 | 96 | 1.30 | 2.17 |
| **2013** | 241 | 20% | 157 | 84 | 1.45 | 2.23 |
| **2014** | 216 | 16% | 139 | 77 | 1.50 | 2.33 |
| **2015** | 240 | 20% | 173 | 67 | 1.55 | 2.16 |
| **2016** | 289 | 17% | 190 | 99 | 1.29 | 1.97 |
| **2017** | 277 | 17% | 196 | 81 | 1.38 | 1.95 |
| **2018** | 288 | 15% | 192 | 96 | 1.24 | 1.86 |
| **Total** | 1792 | 19% | 1192 | 600 | 1.38 | 2.08 |

\*Note: articles without a reference were excluded

**Table 6.2.** Textbooks as references.

|  | **DPRK textbooks** | **DPRK textbooks/all DPRK ref** | **English textbooks** | **English Textbooks/all English ref** | **Chinese textbooks** | **Chinese textbooks/all Chinese ref** |
| --- | --- | --- | --- | --- | --- | --- |
| **2012** | 45 | 71% | 10 | 5% | 0 | 0% |
| **2013** | 59 | 73% | 10 | 5% | 0 | 0% |
| **2014** | 48 | 72% | 21 | 11% | 0 | 0% |
| **2015** | 65 | 69% | 13 | 6% | 0 | 0% |
| **2016** | 70 | 74% | 16 | 7% | 1 | 4% |
| **2017** | 65 | 76% | 33 | 14% | 5 | 12% |
| **2018** | 52 | 76% | 26 | 11% | 0 | 0% |

Most of the original articles were composed of *Gyo-si* (a quote by the Chairman, optional), study population, study method, study results, conclusion, references, and keywords. “Discussion” articles had similar structures but references were not present in this type of article. “Experiences” articles had similar structure that consists study population, methods, results and conclusion without references. Out of 1,792 articles, about 36% had teachings from Kim Jong-il or Kim Jong-un as *Gyo-si*. (Table S1) Kim Jong-un’s teaching first appeared twice in 2018. Contents of these quotes were not directly linked to the topic of the article. Medical images such as X-ray, CT or MRI images were never included in the articles.

### 2. Analysis by disease category

In total, trauma is composed of 25.7% of journal articles that covered surgical diseases. 4.6% of them were about neonatal/pediatric/congenital diseases. Non-communicable diseases were covered in 35.2% of research articles.

### 3. Analysis by specialty

From 2012 to 2018, about 29.6% of articles were written about diseases that undergone orthopedic surgery, and 29.5% were for general surgery. The proportion of cardiothoracic surgery (8.6%), plastic and reconstructive surgery (7.5%), and neurological surgery (6.2%) were followed.

#### General Surgery

General surgery included surgeries on the lower and upper gastrointestinal (GI) tract, hepatobiliary system, vascular system, thyroid, and breast.

##### Trauma

There were 6 reports on trauma cases, 2 on splenic injury, 2 on liver injury, and 2 on traumatic hemorrhagic shock.

##### Pediatric

In the case of pediatric surgery, there were 5 intussusception papers, 4 hernia papers, 3 biliary ectasia papers, 2 megacolon cases, 1 hypertrophic pyloric stenosis and 1 Meckel’s diverticulum case reported from 2012 to 2018.

##### Cancer

For upper GI cases, there were papers on chemotherapy for advanced gastric cancer. Also, some papers were using endoscopic surgery for gastric cancer. There was a paper that specifically covered the topic of MALT lymphoma, which is highly related to *Helicobacter pylori* and size of study population was 2,483. For cholangiocarcinoma, there were 118 cases, 84 were male. The paper states that cholangiocarcinoma is most common in 50s in DPRK. 6 breast cancer surgery reports including 2 on oncoplastic techniques were observed. Also, there was a paper that claimed that mortality of rectal cancer was, for 1996-2005, 1.97% for men and 1.34% for women.

##### Non-communicable diseases

For lower GI cases, there were 29 papers on hemorrhoids, anal fistula, or anal fissure that included anal surgeries. In this period of 2012-2018, 19 appendicitis/appendectomy cases were reported. Surgical treatment of vascular diseases including TAO, ASO, DVT was notable.

##### Others

21 papers included the usage of laparoscopy. Laparoscopic surgeries were used for appendectomy, cholecystectomy, hernia repair (both pediatric and adult), early gastric cancer, and colectomy. No case was reported for laparoscopic lobectomy of liver or laparoscopic surgeries for pancreato-biliary system. There was no paper reporting thyroid surgery for neither benign nor malignant cases.

#### Orthopedic Surgery

##### Trauma

46.7% (248 out of 531) of orthopedic papers were trauma cases which included fractures.

##### Pediatric

For pediatric cases, there were 5 papers on developmental dysplasia of the hip (DDH) and 3 papers on club foot. In 2015, there was a paper with a study population of 2,250 for DDH.

##### Cancer

5 papers were reporting about tumor; one case on chondroma, one on leiomyoma, one on osteoma, and the last on sarcoma. Animal experiments on sarcoma also existed.

##### Non-communicable diseases

Also, 27 papers dealt with back pain including herniated intervertebral disc (HIVD), spinal stenosis, and spondylosis, while 9 papers were on arthropathy or arthritis. 26 papers were on avascular necrosis (AVN), containing both radiological and clinical factors. 7 papers provided cases on synovitis, bursitis, or tendinitis.

##### Others

There were 6 papers using 3D FEM for study, 6 papers on bone substitutes, and 2 papers reporting the cases with arthroscopy. 24.7% (131 out of 531) of the orthopedic papers utilized X-ray in their studies; 4.5% (24 out of 531) and 3.2% (17 out of 531) used CT and MRI respectively (with and without contrast). Novocaine (procaine) was used in 19 papers and the usage of lidocaine was found in 11 papers.

#### Cardiothoracic Surgery

##### Trauma

There were 6 papers on trauma.

##### Pediatric

For pediatric cases, there were 3 papers on Tetralogy of Fallot, 1 case report on Marfan syndrome, 6 papers on ventricular septal defect (VSD), and 4 papers on patent ductus arteriosus (PDA).

##### Cancer

There were 12 papers on lung cancer and 1 paper on a mediastinal tumor. One author (S.R., Ahn) was included in 25% of the paper on lung cancer (3 out of 12). While lung adenocarcinoma is distinguished into tubular, papillary, mucinous and poorly cohesive type pathologically according to WHO 2010 classification, the 2015 paper that studied survival rate of lung adenocarcinoma classified it into mucinous and non-mucinous type.

##### Non-communicable diseases

13 papers handled the problem of valves; studies on mitral valve accounted for 76.9% (10 out of 13). 2 papers were on pulmonary valve, and the other on aortic valvular disease. There were 3 papers examining the treatment of pleuritis, 1 paper on pericarditis, and 2 papers on pleural effusion. One study reported the case of cardiac tamponade. There were also 3 papers on pneumothorax. 2 papers on thoracic sympathectomy and 4 papers on coronary artery bypass grafting (CABG) were also identified.

##### Others

24.7% (38 out of 154) and 4.5% (7 out of 154) of the cardiothoracic papers used X-ray and CT respectively. 23.4% (36 out of 154) utilized ultrasonography. There were 6 papers that used dimedrol, 5 papers using morphine, and 4 papers using novocaine (procaine) for anesthesia.

#### Neurological Surgery

##### Trauma

For other cases like epidural hemorrhage (EDH), subdural hemorrhage (SDH), intracerebral hemorrhage (ICH) except for the skull fracture, incidences in men were higher. There were 2 papers on EDH, 9 papers on SDH, and 3 papers on ICH. One reported the treatment of parkinsonism caused by CO poisoning.

##### Pediatric

There were 4 congenital disease papers – 3 on moyamoya disease, 1 on cerebral palsy. There was no paper on pediatric hydrocephalus. But 3 papers on adult hydrocephalus. All hydrocephalus papers used ETV as surgical method, except for one paper describing the characteristics of normal pressure hydrocephalus (NPH).

##### Cancer

There were 11 papers on brain or spine tumors from 2012 to 2018. For brain tumors, there were 2 papers on CPA tumor, 2 papers on meningioma, and 2 papers on the pituitary tumor. In these papers, pituitary tumors were approached using trans-sphenoidal approach (TSA).

##### Non-communicable diseases

2 papers were on myelitis, while 1 paper reviewed the features of ischemic strokes.

##### Others

25.2% (28 out of 111) of the neurological papers were using CT, the usage of MRI accounted for 9.9% (11 out of 111). 5 papers were using novocaine (procaine).

#### Plastic and Reconstructive Surgery

Burn and wound were common topics. There were 51 articles (35.2%) covering the topic of burn, and 32 articles (22.1%) on the topic of a wound. Especially for the treatment of burn, papers reporting the usage of oriental/herbal or Koryo medicines were noticed. Koryo Burn Ointment, arsenic sulfide, extraction from lilac leaves, egg white – rifampicin mix, *Coptis chinensis, Cirsium japonicum var. maackii, Lonicera japonica* are the examples. A case report with treatment of electric injury existed. The skin flap was also highlighted with 26 articles (17.9%); 4 papers on amputation, 3 papers on frostbite, 1 report on scalp laceration, and 3 papers on pressure sore were reported.

There were 2 congenital disease papers; case reports of syndactyly and microtia were provided. Only 2 papers utilized X-ray, while a single article examined the clinical usage of pre-operational doppler sonography on skin flap. 9 papers reported the usage of novocaine (procaine).

#### Urology

##### Trauma

There were 8 trauma papers and among those five were on traumatic urethral stricture

##### Pediatric

There were 6 pediatric papers on CAH, circumcision, phimosis, cryptorchidism, Wilm’s tumor, hypospadias.

##### Cancer

There were 12 papers on urologic cancers. Bladder cancer accounted for half of the articles; prostate cancer, cystic cancer, and renal cancer were also included.

##### Non-communicable diseases

There were 5 papers on benign prostate hyperplasia (BPH), 1 paper on sparganosis, and 10 papers covering stones in the urinary tract.

##### Others

10 papers utilized ultrasonography in the studies; a single paper reported the usage of cystoscopy. 3 papers used novocaine (procaine), including a case report to treat idiopathic priapism with epidural anesthesia.

### 4. Other characteristics

#### 1) Reports on oriental medicine

About 3.3% of the papers utilized oriental medicine (Koryo Medicine) for their research; oriental medicine may include various types of treatment such as herbal therapy, acupuncture, physical exercise, or diet. (https://www.cancer.gov/publications/dictionaries/cancer-terms/def/oriental-medicine, accessed October 24^th^, 2020.) The proportion of reference on oriental medicine varied with the published periods from 1.7% in 2015 to 4.6% in 2012 but remained under 5% in all periods.

#### 2) Study population

Besides researches on the human population, studies using cadaver or animals were reported. Average 2% of articles used cadavers and 6% used animal models.

### 5. Analysis of references

Around 33% of articles were without reference, and the average number of references was 1.38. Out of all references cited, about 20% of them were references published in recent 5 years. Average 22.3% of references were North Korean journals or textbooks. 12.8% of references were in Chinese, 2.7% were in Japanese and 60.7% were in English. From 2012 to 2018, there were 9 references (0.4%) in German and 15 references (0.6%) in Russian. Textbooks were composed of 76% of all North Korean references. 11% of English references were textbooks.

### 5. Imaging and anesthesia

The utilization of medical imaging techniques such as radiography could be found in the papers. The trends of usage in medical devices from 2012 to 2018 are shown in Table 7. About 11.9% of the researchers utilized X-ray, while the usage of CT or MRI remained under 5% (4.4% and 1.8% each). Ultrasound was the second commonly utilized medical imaging technique listed in the journals (9.0%). Articles utilizing single imaging devices accounted for 20.3% while articles with multiple devices accounted for 5.9%. 73.8% of papers did not take advantage of the imaging techniques. For imaging devices, some articles introduced them with specific model names. For instance, EDR-750b, *Bulgeungi* 66(Russian product), F-30III, *Tamjeong* were used. For endoscopy, Olympus gif-d2, Intravision-8745, Model MGA-III were used and for ultrasound, Aloka ssd-650: Aloka-SSD 620 marketed 1988 discontinued 1996, HP SONOS 5500, 2.5MHz marketed 1988, ATL Sono-CT HDI-5000. 1996 – 2006, Medison 128BW, Medison SonoAce 8000, Sonoace 8800 were used. (cannot be specified as these are manufactured for a long time with different editions) Aloka SSD-650 was marketed in 1986 and discontinued in 1997 (http://www.ob-ultrasound.net/manufacturers.html). In 2014, 2013, 2012, there is no specific imaging device described.

**Table 7.** Medical imaging techniques utilized in North Korean surgery journals.

|  | **X-ray** | **CT<sup>†</sup>** | **MRI<sup>†</sup>** | **Endoscopy** | **Fluoroscopy** | **USS** | **Angiography** |
| --- | --- | --- | --- | --- | --- | --- | --- |
| 2012 | 22<br>(9.1%) | 11<br>(4.6%) | 7<br>(2.9%) | 3<br>(1.2%) | 7<br>(2.9%) | 18<br>(7.5%) | 2<br>(0.8%) |
| 2013 | 33<br>(13.7%) | 13<br>(5.4%) | 7<br>(2.9%) | 2<br>(0.8%) | 6<br>(2.5%) | 31<br>(12.9%) | 4<br>(1.7%) |
| 2014 | 28<br>(13.0%) | 5<br>(2.3%) | 2<br>(0.9%) | 6<br>(2.8%) | 4<br>(1.9%) | 15<br>(6.9%) | 1<br>(0.5%) |
| 2015 | 21<br>(8.8%) | 8<br>(3.3%) | 5<br>(2.1%) | 9<br>(3.8%) | 12<br>(5.0%) | 20<br>(8.3%) | 0<br>(0.0%) |
| 2016 | 35<br>(12.1%) | 14<br>(4.8%) | 2<br>(0.7%) | 7<br>(2.4%) | 11<br>(3.8%) | 31<br>(10.7%) | 1<br>(0.3%) |
| 2017 | 34<br>(12.3%) | 12<br>(4.3%) | 4<br>(1.4%) | 7<br>(2.5%) | 7<br>(2.5%) | 21<br>(7.6%) | 4<br>(1.4%) |
| 2018 | 41<br>(14.2%) | 15<br>(5.2%) | 6<br>(2.1%) | 4<br>(1.4%) | 12<br>(4.2%) | 25<br>(8.7%) | 5<br>(1.7%) |
| Total | 214<br>(11.9%) | 78<br>(4.4%) | 33<br>(1.8%) | 38<br>(2.1%) | 59<br>(3.3%) | 161<br>(9.0%) | 17<br>(0.9%) |

Note. <sup>†</sup>CT and MRI include both cases with or without contrast. CT = Computed Tomography. MRI = Magnetic Resonance Imaging. USS = Ultrasound Scan.

**Table 8.** Commonly used anesthetics and supplementary medications.

|  | Novocaine | Dimedrol | Morphine | Lidocaine | Fentanyl | Atropine | Ketamine | Thiopental | Diazepam |
| --- | --- | --- | --- | --- | --- | --- | --- | --- | --- |
| 2012 | 15<br>(6.2%) | 9<br>(3.7%) | 11<br>(4.6%) | 8<br>(3.3%) | 4<br>(1.7%) | 4<br>(1.7%) | 1<br>(0.4%) | 4<br>(1.7%) | 4<br>(1.7%) |
| 2013 | 14<br>(5.8%) | 4<br>(1.7%) | 5<br>(2.1%) | 5<br>(2.1%) | 4<br>(1.7%) | 3<br>(1.2%) | 2<br>(0.8%) | 6<br>(2.5%) | 2<br>(0.8%) |
| 2014 | 14<br>(6.5%) | 7<br>(3.2%) | 7<br>(3.2%) | 11<br>(5.1%) | 3<br>(1.4%) | 3<br>(1.4%) | 5<br>(2.3%) | 3<br>(1.4%) | 2<br>(0.9%) |
| 2015 | 5<br>(2.1%) | 10<br>(4.2%) | 7<br>(2.9%) | 8<br>(3.3%) | 10<br>(4.2%) | 7<br>(2.9%) | 9<br>(3.8%) | 3<br>(1.3%) | 4<br>(1.7%) |
| 2016 | 15<br>(5.2%) | 2<br>(0.7%) | 3<br>(1.0%) | 6<br>(2.1%) | 5<br>(1.7%) | 2<br>(0.7%) | 4<br>(1.4%) | 4<br>(1.4%) | 2<br>(0.7%) |
| 2017 | 9<br>(3.2%) | 6<br>(2.2%) | 6<br>(2.2%) | 4<br>(1.4%) | 3<br>(1.1%) | 7<br>(2.5%) | 3<br>(1.1%) | 2<br>(0.7%) | 5<br>(1.8%) |
| 2018 | 8<br>(2.8%) | 10<br>(3.5%) | 8<br>(2.8%) | 3<br>(1.0%) | 6<br>(2.1%) | 7<br>(2.4%) | 2<br>(0.7%) | 3<br>(1.0%) | 2<br>(0.7%) |
| Total | 80<br>(4.5%) | 48<br>(2.7%) | 47<br>(2.6%) | 45<br>(2.5%) | 35<br>(2.0%) | 33<br>(1.8%) | 26<br>(1.5%) | 25<br>(1.4%) | 21<br>(1.2%) |

**Table 9.** Essential surgeries covered in the Journal ‘Surgery’.

| **Essential Surgery** | **Covered by North Korean Surgery Journal** |
| --- | --- |
| 1. Drainage of superficial abscess | X |
| 2. Male circumcision | O |
| 3. Repair of perforation | O |
| 4. Appendectomy | O |
| 5. Bowel obstruction | O |
| 6. Colostomy | X |
| 7. Gallbladder disease | O |
| 8. Hernia | O |
| 9. Hydrocelectomy | O |
| 10. Relief of urinary obstruction; suprapubic cystostomy | O (relief of urinary obstruction) X (suprapubic cystostomy) |
| 11. Suturing laceration | O |
| 12. Management of non-displaced fractures | O |
| 13. Tube thoracostomy | O |
| 14. Trauma laparotomy | O |
| 15. Fracture reduction | O |
| 16. Irrigation and debridement of open fractures | O |
| 17. Placement of external fixator; use of traction | O |
| 18. Escharotomy or fasciotomy | X |
| 19. Trauma-related amputations | O |
| 20. Skin grafting | O |
| 21. Burr hole | O |
| 22. Cleft lip and palate repair | X |
| 23. Club foot repair | O |
| 24. Shunt for hydrocephalus | X (only ETVs were mentioned) |
| 25. Repair of anorectal malformation | X |
| 26. Hirschsprung's disease | X |
| 27. Drainage of septic arthritis | O |
| 28. Debridement of osteomyelitis | O |

Various kinds of injection materials were utilized for anesthesia; 56 types of medications were listed to be utilized in the articles. Commonly used anesthetics and supplementary medications are listed in Table 2. Novocaine (Procaine) was the most commonly exploited anesthetics (4.5%). Papers specifying their usage of medications accounted for 11.6% of overall publications.

### 6. Research Methodology

The trends of specific research methodology are shown in Figure 1. The proportion of case-control studies increased more than 10%p, from 29.9% in 2012 to 40.3% in 2018. The proportion of observational study and case report (or series) dramatically decreased (29.5% to 19.4%; 27.0% to 16.3%). The statistics stressed the increasing attention on basic science; the number of papers on basic science surged in recent years. Editorial papers on their ideology were consistently published in the beginning of the journals.

**Figure 1.**
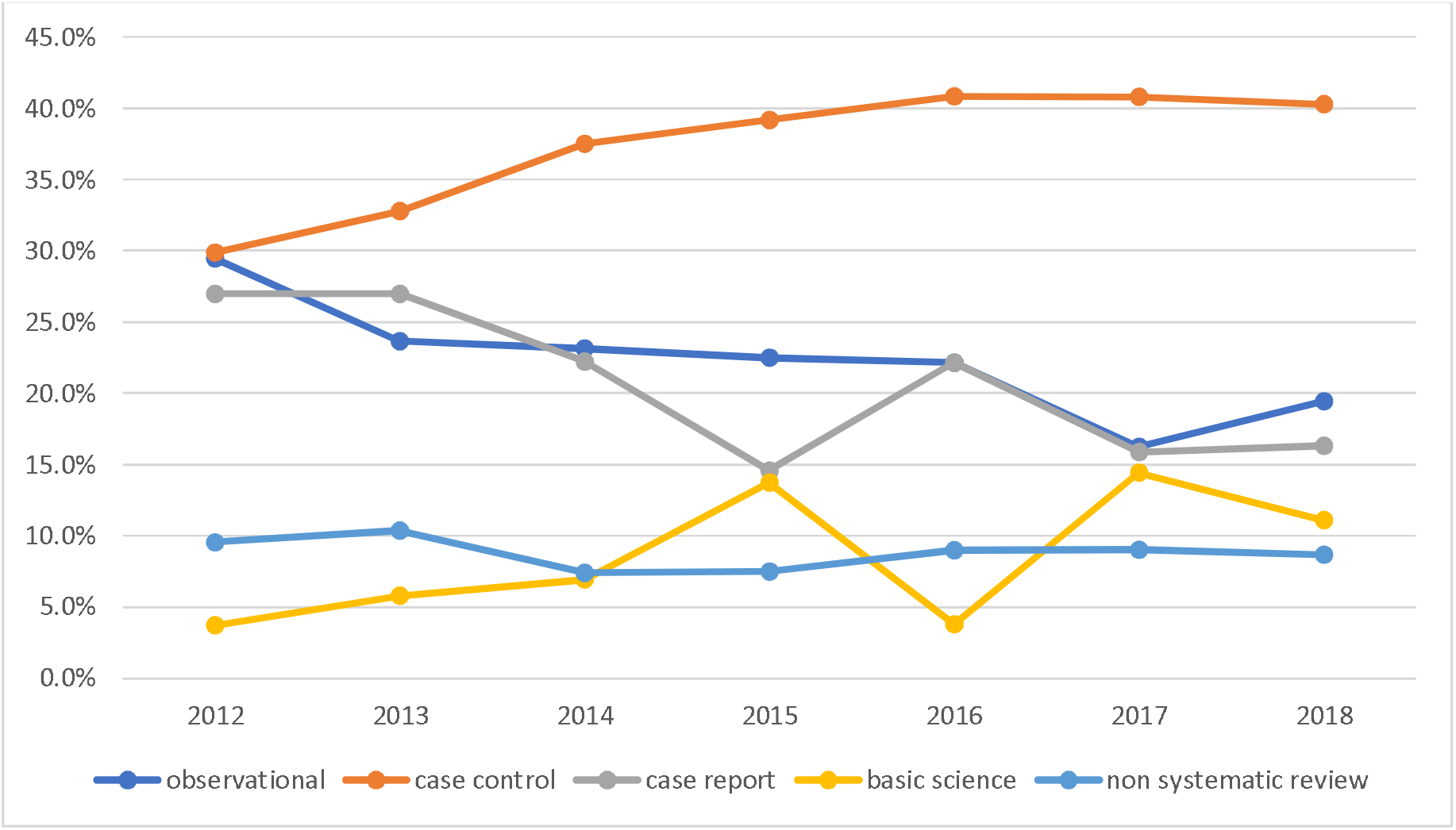
Trends of research methodology in North Korean surgery journals.

## DISCUSSION

This study showed the trends of researches performed in North Korea in the surgical field during the Kim Jong-un era. Analysis by specialty and disease category reflected status and characteristics of surgical diseases in North Korea.

### Surgical diseases in Kim Jong-un Era

Significant changes have been made in the sector of health and medicine since the beginning of Kim Jong-un era. (Shin and Jeon 2019) The establishment of hospitals, medical research institutes, and factories for medical supplies are contributing to the development of active research outcomes; the authorities are attempting to renovate and modernize the facilities. (Shin, Lee et al. 2016) While centralized facilities appear to be the limiting point for improving the system, general advancement of the field is affecting the quality of clinical infrastructure in surgery as a result. Specific plans were established to develop medical instruments and introduce up-to-date study outcomes to each clinical field; the medical associations have even newly organized a committee for plastic surgery in 2020. Detailed results of the efforts are specifically illustrated in the North Korean journal ‘Surgery’.

#### Structure

Articles published in the journal were relatively short in length - on average ½ to 1 page per paper. It is possible that the papers attached in the journal are not full papers but abstracts.

#### Imaging & Anesthesia

For imaging, there were 78 papers (4.4%) that mentioned CT in the study method and 33 (1.8%) papers with MRI as imaging devices. Considering this proportion, CT and MRI may not be widely used imaging modalities in North Korea.

#### Disease Prevalence and Burden

The high proportion of topics related to non-communicable diseases in the journal ‘Surgery’ (35.2%) supports the general prevalence in North Korea. Non-communicable diseases including tumors account for eight among top 10 causes of total number of deaths in North Korea (Available from http://www.healthdata.org/north-korea, Accessed 25-10-2020); lung cancer and stomach cancer are fourth and sixth common cause of deaths. The overall trends of South Korea do not differ much from the statistics of North Korea; lung cancer, liver cancer, stomach cancer, and colorectal cancer are investigated to be the third, seventh, ninth, and tenth common causes respectively (Available from http://www.healthdata.org/north-korea, Accessed 25-10-2020). The academic interest in non-communicable diseases might be much larger in North Korea considering the articles in the other journals not included in the analysis of the study.

However, the percentage of articles on the topic of neoplasm among non-communicable diseases were relatively low compared to the actual prevalence. Some types of cancers such as thyroid cancer that were reported to be frequent in North Korean were not even handled (Park, Kim et al. 2014) The steady publications of articles on various types of cancers implies continuous participation and interest in the field, but the lack of healthcare facilities capable of cancer management may have resulted in the shortage of research outcomes. There was barely any glioma case, but pituitary tumors and trans-sphenoidal approach were published. According to Park’s paper (Park, Roh et al. 2015), WFNS delivered Zeiss Pico Microscope in 2010, so it’s not solely a matter of equipment.

Besides, Certain surgical treatments such as liver transplantation, kidney transplantation were not covered. From this, it can be assumed that organ transplantation is not or barely performed in North Korea. This could be because of lack of surgical technique or lack of pre-/post-op care which requires intensive care facilities.

#### Trauma

There was a high percentage of trauma/injury cases – ranging from traumatic brain injury accompanied by hemorrhages, fractures, liver, or splenic injuries.

#### Hospital

Since the authors’ affiliation is not written in the paper, and there was no location information, it is difficult to speculate where these patients were collected from. Considering substandard conditions of provincial hospitals or local clinics in North Korea, it is reasonable to assume that these papers were based on tertiary hospitals in Pyongyang. (Shin, Lee et al. 2016)

#### Medical Device

Due to sanction and political situation, in DPRK, they try to develop their medical equipment including essential spinal implants (Park, Roh et al. 2015), spiral CT from Kim Chaek University (2016 issue), and prosthetic legs made out of unsaturated resin from Ham Heung (2016 issue)

#### Medical conference

In 2017, in issue 1 of Surgery journal, it is stated that the 23^rd^ National Cardiothoracic Conference, 34^th^ National Abdominal Surgery Conference, and 4^th^ Resuscitation Conference were held. It can be assumed that regular medical conferences by specialty are being held in DPRK.

### In the Context of Global Surgery (Mock, Donkor et al. 2015, Sullivan, Alatise et al. 2015, Ng-Kamstra, Greenberg et al. 2016)

World Health Organization suggested 44 essential surgeries in 2015 to reduce the burden and surgically avertable deaths worldwide. Out of 44 essential surgeries, besides ophthalmology, obstetrics and gynecology, and dental surgeries, 28 were selected for comparison with researches in North Korean Journal ‘Surgery’. 8 of them could not be found - drainage of superficial abscess, suprapubic cystostomy, or escharotomy can be too simple and basic to be a research topic. However, it is important to notice that others that were not covered were congenital diseases like cleft lip, anorectal malformation, and Hirschsprung’s disease. Needs for surgeries on congenital diseases in North Korea should be investigated further.

### Limitations

This study has several limitations. First, this is the study with an indirect approach to estimate the epidemiology of DPRK’s surgical needs as we used published journals as sources of analysis. Research papers reflect the research interests of North Korean doctors and not exactly the disease characteristics or burdens of North Korea. Also, these research papers did not indicate where their study population resides, which could be led to disproportionate geographical distribution of study population. A direct survey of the North Korean population, which is almost impossible in current political situation, is the way to collect unbiased and reliable data. Second, the scope of the study only included the Jong-un Kim period. Data of previous periods could have given us more whole picture.

## Data Availability

Data is available at North Korea Resource Center in Seoul, South Korea when requested. There is no digital format of the data.

## Funding Acknowledgement

Seoul National University College of Medicine

## SUPPLEMENTARY

**Table S1.**
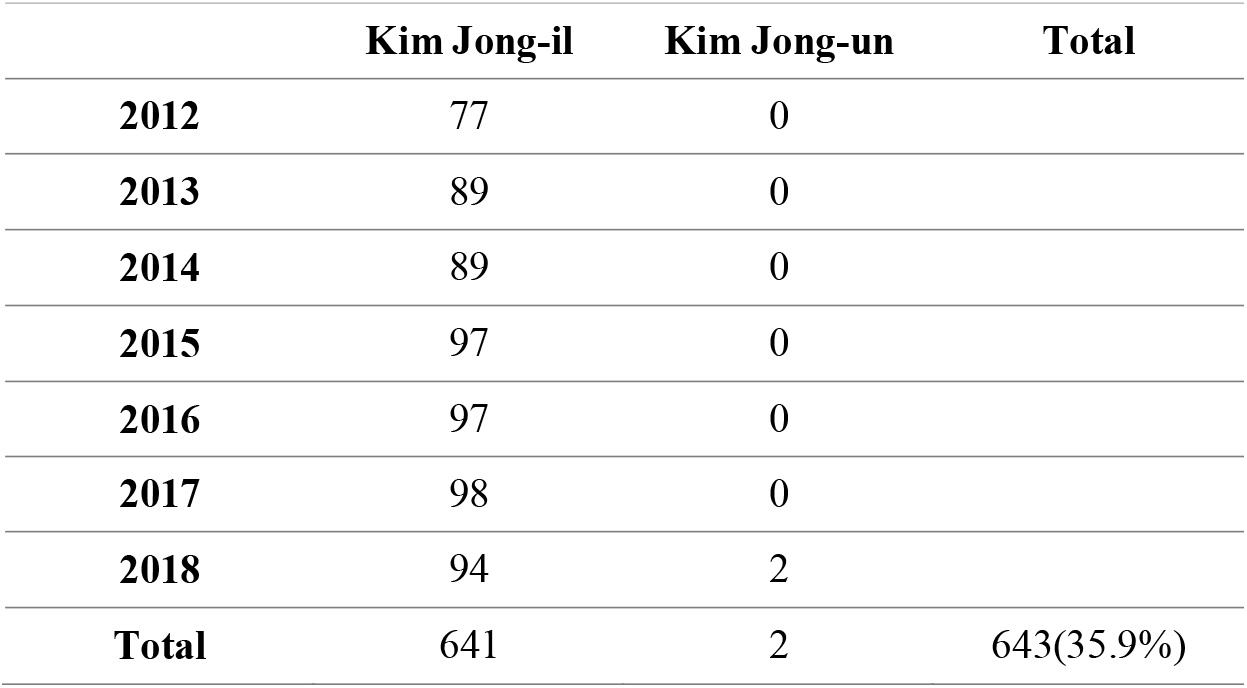
Gyo-si by chairman of DPRK.

